# Trends of COVID-19 (Coronavirus Disease) in GCC Countries using SEIR-PAD Dynamic Model

**DOI:** 10.1101/2020.11.29.20240515

**Authors:** Ahmad Sedaghat, Seyed Amir Abbas Oloomi, Mahdi Ashtian Malayer, Amir Mosavi

## Abstract

Extension of SIR type models has been reported in a number of publications in mathematics community. But little is done on validation of these models to fit adequately with multiple clinical data of an infectious disease. In this paper, we introduce SEIR-PAD model to assess susceptible, exposed, infected, recovered, super-spreader, asymptomatic infected, and deceased populations. SEIR-PAD model consists of 7-set of ordinary differential equations with 8 unknown coefficients which are solved numerically in MATLAB using an optimization algorithm. Four set of COVID-19 clinical data consist of cumulative populations of infected, deceased, recovered, and susceptible are used from start of the outbreak until 23^rd^ June 2020 to fit with SEIR-PAD model results. Results for trends of COVID-19 in GCC countries indicate that the disease may be terminated after 200 to 300 days from start of the outbreak depends on current measures and policies. SEIR-PAD model provides a robust and strong tool to predict trends of COVID-19 for better management and/or foreseeing effects of certain enforcing laws by governments, health organizations or policy makers.

## Introduction

The coronavirus was first observed in chickens in 1930 and no traces were observed in humans until 1960. Seven type of coronaviruses to date were recognised which affected humans. Four types of the coronaviruses were not fatal. This include the coronavirus type 229E and OC43 which caused common cold least severe disease; the coronavirus NL63 which caused suffering for babies from bronchiolitis in the Netherlands in 2004; and the coronavirus HKU1 which affected elderlies with pneumonia in Hong-Kong in 2005 [1].

In 2002, the fatal coronavirus with severe acute respiratory syndrome (Sars) named SARS-CoV was first observed in humans which is very similar to the current COVID-19. This virus affected elderly people with symptoms included fever, sore throat, cough, and muscle pain from 2002 until 2014. A deadlier coronavirus appeared after nearly a decade in Saudi Arabia known as Middle East respiratory syndrome coronavirus (MERS-CoV) in 2012 [2].

The MERS-CoV had largest impact in Saudi Arabia and reappeared in 2015 in South Korea and in 2018 in Saudi Arabia and United Arab Emirates which caused more than 35% mortality rate among infected people with fever, cough, and shortness of breath [3, 4]. Alasmawi *et al*. [5] studied MERS-CoV by an extended SEIR model consist of 5 set of population included susceptible (S), infected (I), super-spreaders (P), recovered (R), and hospitalized (H). The model coefficients were adopted from previous literature and focus of study was on determining the re-production number (R_0_). Kim *et al*. [6] studied outbreak of MERS-CoV in South Korea using SEIR model of Alasmawi *et al*. with addition of asymptomatic infected population (A). They only use one set of clinical data on cumulative incidence cases numbers to suggest super spreader population influences on high initial reproduction number.

Ndaïrou *et al*. [7] applied similar SEIR type model for studying outbreak of COVID-19 in Wuhan, China with addition of fatality population in the model. Two set of clinical data including confirmed daily cases and daily death were used.

Recently, Xue *et al*. [8] used similar SEIR type model for COVID-19 in Wuhan (China), Toronto (Canada), and the Italy. They found model coefficients using the optimization algorithm (MCMC) yet they validated their model against only two set of clinical data.

In the above literature, it is ambiguous why 5 to 8 set of ordinary differential equations (ODE) should be used to study only one-parameter or one or two set of clinical data. With recent COVID-19 development, we can easily use 4 set of clinical data for validation of any SEIR type models. In this paper, we have developed SEIR-PAD model compose of 7 populations influenced by COVID-19 outbreak. The aim is to computationally predict trends of COVID-19 in GCC countries. SEIR-PAD model ODE equations are solved using an optimization technique (fminsearch) in MATLAB to find best model coefficients utilizing available clinical data in GCC countries.

## Methodology

### SEIR-PAD Model

SEIR-PAD model developed here consist of 7 populations starts, similar to original SIS and SIR model reported by Kermack and McKendrick [9], with a different nonlinear transmission rate 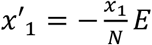 from susceptible population; as shown in the flow chart of the model in Figure 1.

**Figure 1:**
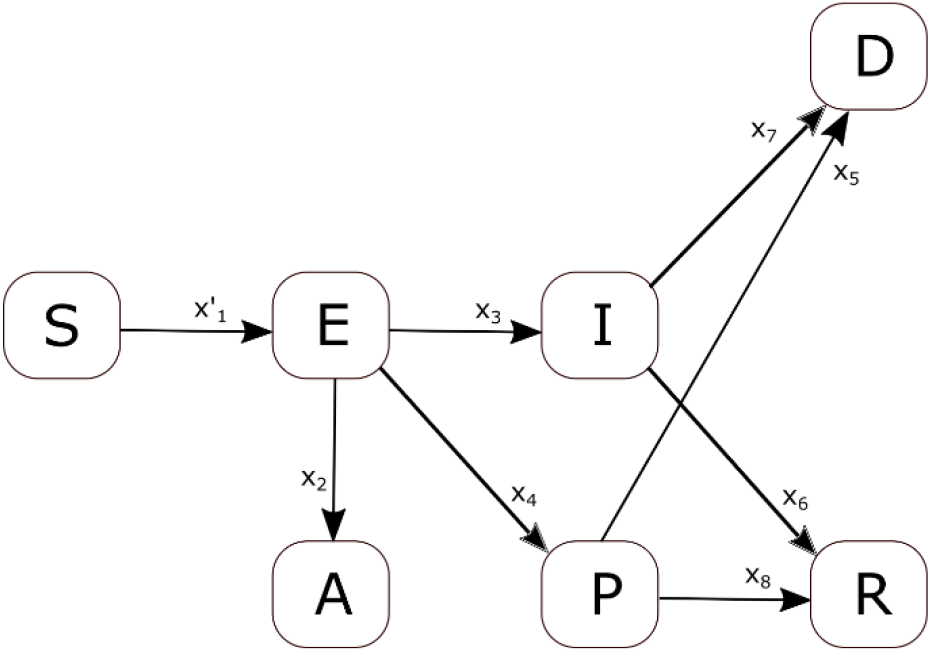
Flow chart of SEIR-PAD model.

SEIR-PAD model has 8 unknown transmission coefficients x=[x_1_ x_2_ …x_8_] that can be obtained by an optimization algorithm in MATLAB by fitting available clinical data. The rest of transmission coefficients simply linearly relate flow in and flow out of populations as follows:

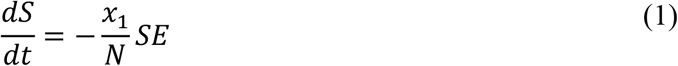

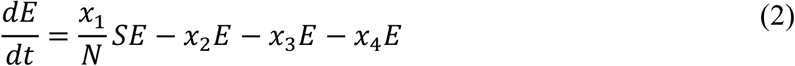

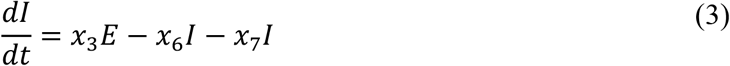

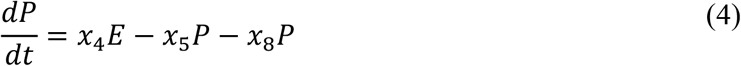

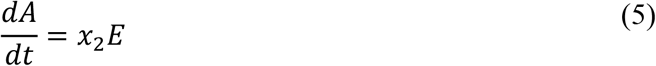

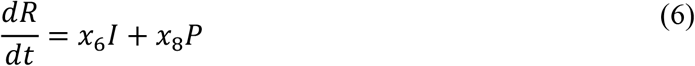

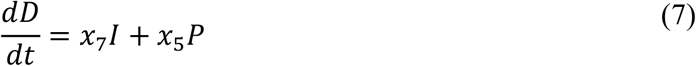

The total population *N* is constant and is defined by:

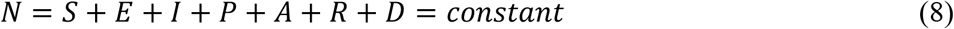

GCC countries consist of Bahrain with 1,701,575, Kuwait with 4,270,571, Oman with 5,106,626, Qatar with 2,881,053, KSA with 34,813,871, and UAE with 9,890,402 populations based on Worldometer in 2020 [10]. To solve SEIR-PAD model 7 ODEs (1-7), one set of initial conditions for populations and the following initial conditions for transmission rate coefficients are applied:

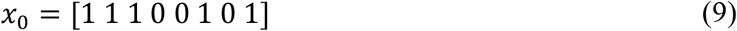

MATLAB ode45 solver [11] is used together with the optimization tool (fminsearch) [12] to find best fitted solutions to the available COVID-19 clinical data using the following convergence criterion:

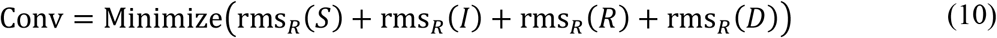

Equation (10) is used for minimizing the root-mean-square ratio (rms_*R*_) of the four populations. Fminsearch in MATLAB returns the transmission rate coefficients that minimize the summation in Equation (10).

### Evaluation method

#### Root-mean-square ratio

Root-mean-square ratio (rms_*R*_) uses the coefficient of determination which is broadly used for comparing prediction variables with actual clinical data. The coefficient of determination (R^2^) compares a predicted value (*z*) against clinical data (*z*_*exp*_) to provide rms_*R*_ as follows [13]:

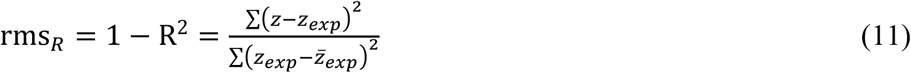

In Equation (11), *z* represents a population and 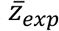 is an average value of the same population. Better prediction yields R^2^ value close to one whilst rms_*R*_ merges to zero.

## Results and Discussions

Table 1 summarizes the used initial conditions for populations and outcome of the optimization algorithm (fminsearch) in MATLAB for the transmission rate coefficients x=[x_1_ x_2_ …x_8_] in Equations (1-7) for different GCC countries.

**Table 1:** Results of SEIR-PAD model for GCC countries

|  |  |
| --- | --- |
| Kuwait | $S_0 = 800,000; E_0 = 3; I_0 = 0; P_0 = 3; A_0 = 0; R_0 = 0; D_0 = 0$ |
| | $x = [1.5039 \ 0.3744 \ 1.0519 \ 0.0043 \ 0.0017 \ 0.0690 \ 0.0007 \ 0.3103]$ |
| Bahrain | $S_0 = 800,000; E_0 = 1; I_0 = 0; P_0 = 1; A_0 = 0; R_0 = 0; D_0 = 0$<br>$x = [2.3442 \ 0.8998 \ 1.3735 \ -0.0008 \ 0.0018 \ 0.0796 \ 0.0002 \ 0.6185]$ |
| Oman | $S_0 = 800,000; E_0 = 2; I_0 = 0; P_0 = 2; A_0 = 0; R_0 = 0; D_0 = 0$<br>$x = [1.1956 \ 0.1840 \ 0.9469 \ 0.0011 \ 0.0077 \ 0.0363 \ 0.0004 \ 1.3131]$ |
| Qatar | $S_0 = 1,400,000; E_0 = 1; I_0 = 0; P_0 = 1; A_0 = 0; R_0 = 0; D_0 = 0$<br>$x = [1.7894 \ 0.9925 \ 0.6735 \ 0.0007 \ 0.0013 \ 0.0494 \ 0.0001 \ 1.2830]$ |
| KSA | $S_0 = 1,600,000; E_0 = 1; I_0 = 0; P_0 = 1; A_0 = 0; R_0 = 0; D_0 = 0$<br>$x = [1.4063 \ 0.6036 \ 0.6872 \ -0.0029 \ -0.0032 \ 0.0805 \ 0.0009 \ 2.0579]$ |
| UAE | $S_0 = 600,000; E_0 = 1; I_0 = 1; P_0 = 0; A_0 = 0; R_0 = 0; D_0 = 0$<br>$x = [1.2121 \ 0.5513 \ 0.5765 \ 0.0011 \ 0.0008 \ 0.0348 \ 0.0005 \ 1.8702]$ |

Figure 2 compares results of SEIR-PAD model for infected, deceased, recovered, and susceptible populations against COVID-19 clinical data available online on 23 June 2020 on Worlometer [10] for Kuwait. Peak day of infection is predicted 13^th^ June 2020 with 12,547 active cases whilst actual peak day was 31 May 2020 with 14,839 active cases (see Fig. 2a). Total expected deceased cases are 567 (see Fig. 2b) cases by expected end of the pandemic after 220 days from start of outbreak; i.e. 1^st^ October 2020. This is based on present policies and preventive measure set for the country. As shown in Fig. 2c, expected total recovered cases by the end of pandemic is predicted 56,100 cases. Total susceptible population will be 723,250 by the end of pandemic (see Fig. 2d).

**Figure 2:**
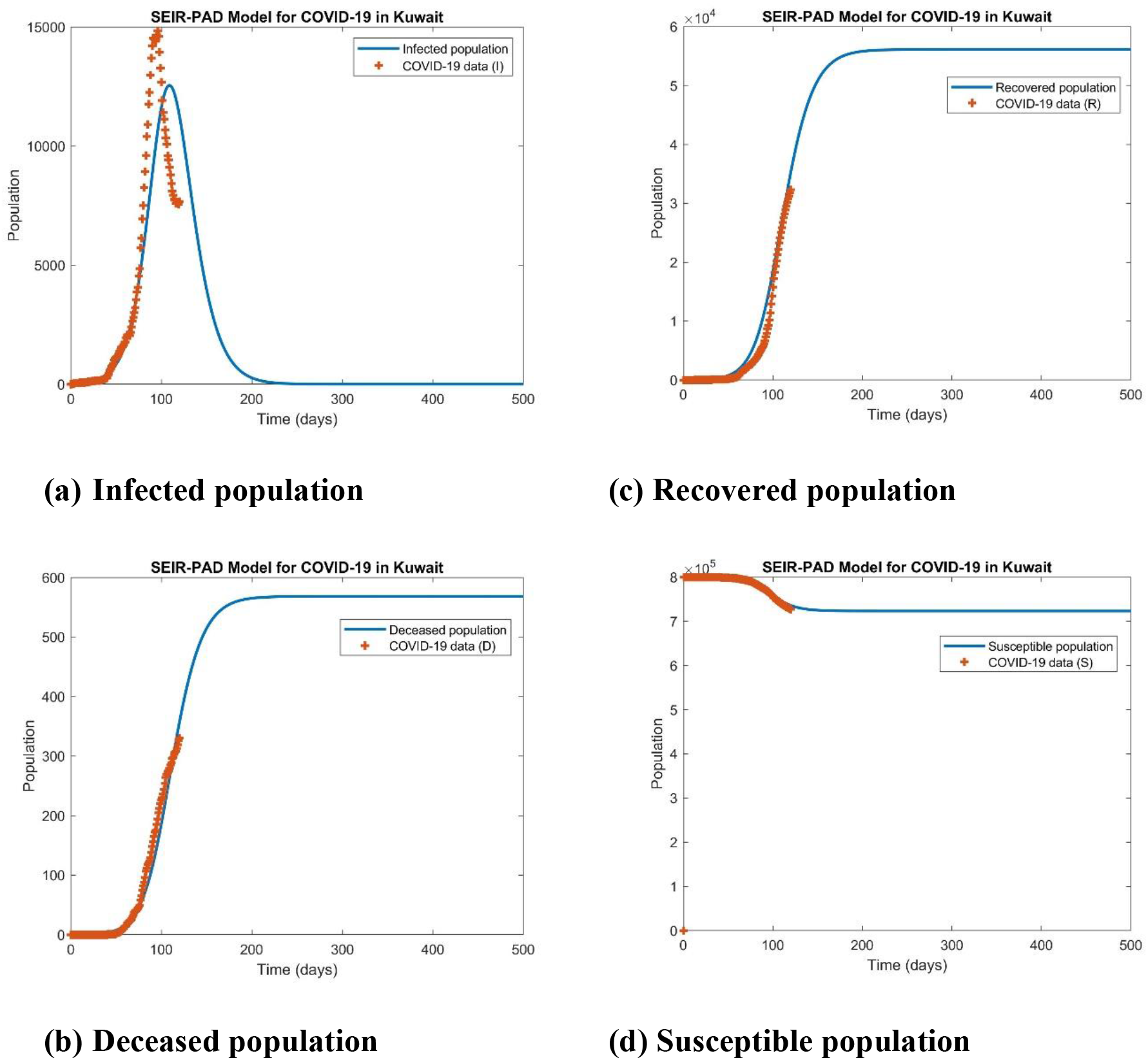
Fitting 4 set of COVID-19 data simultaneously with SEIR-PAD model for dynamics of COVID-19 in Kuwait (23 June 2020).

Figure 3 shows results of SEIR-PAD model for infected, deceased, recovered, and susceptible populations against COVID-19 clinical data on 23 June 2020 from [10] for Bahrain. Peak day of infection is predicted 16^th^ June 2020 with 5,766 active cases whilst actual peak day was 14^th^ June 2020 with 5,535 active cases (see Fig. 3a). Total expected deceased cases are 73 (see Fig. 3b) cases by expected end of the pandemic after 200 days from start of outbreak; i.e. 11^th^ September 2020. This is based on present preventive measures set by the Bahrain government. As shown in Fig. 3c, expected total recovered cases by the end of pandemic is predicted 29,200 cases. Total susceptible population will be 751,540 by the end of pandemic (see Fig. 3d).

**Figure 3:**
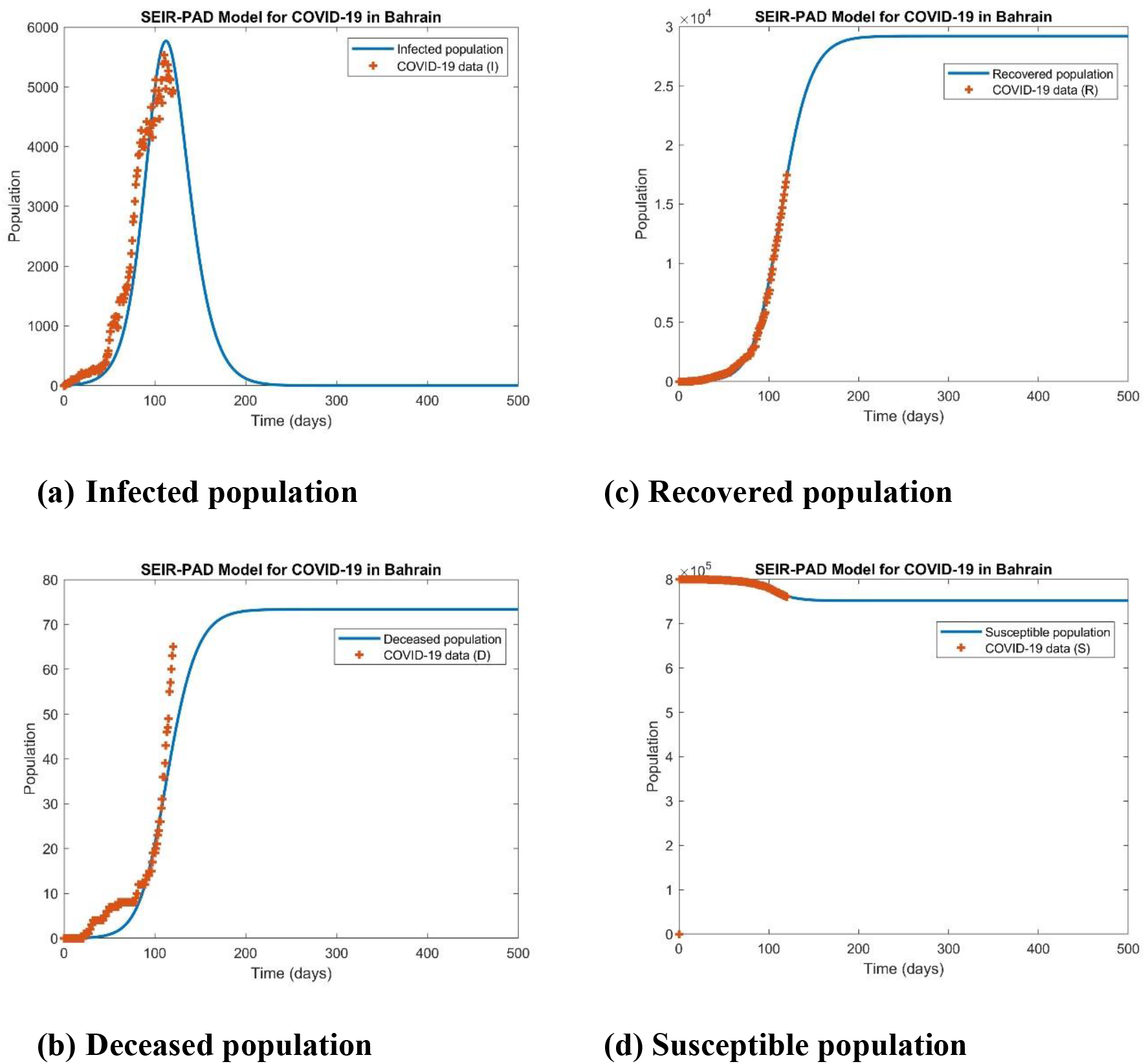
Fitting 4 set of COVID-19 data simultaneously with SEIR-PAD model for dynamics of COVID-19 in Bahrain (23 June 2020).

Figure 4 shows the same results of SEIR-PAD model for Oman. Peak day of infection is predicted 15^th^ July 2020 with 22,357 active cases whilst actual peak day was not yet reached on 23^rd^ June 2020 (see Fig. 4a). Total expected deceased cases are 760 (see Fig. 4b) cases by expected end of the pandemic after 300 days from start of outbreak; i.e. 21^st^ December 2020. This is based on present preventive measures set by the Oman government. As shown in Fig. 4c, expected total recovered cases by the end of pandemic is predicted 69,260 cases. Total susceptible population will be 716,390 by the end of pandemic (see Fig. 4d).

**Figure 4:**
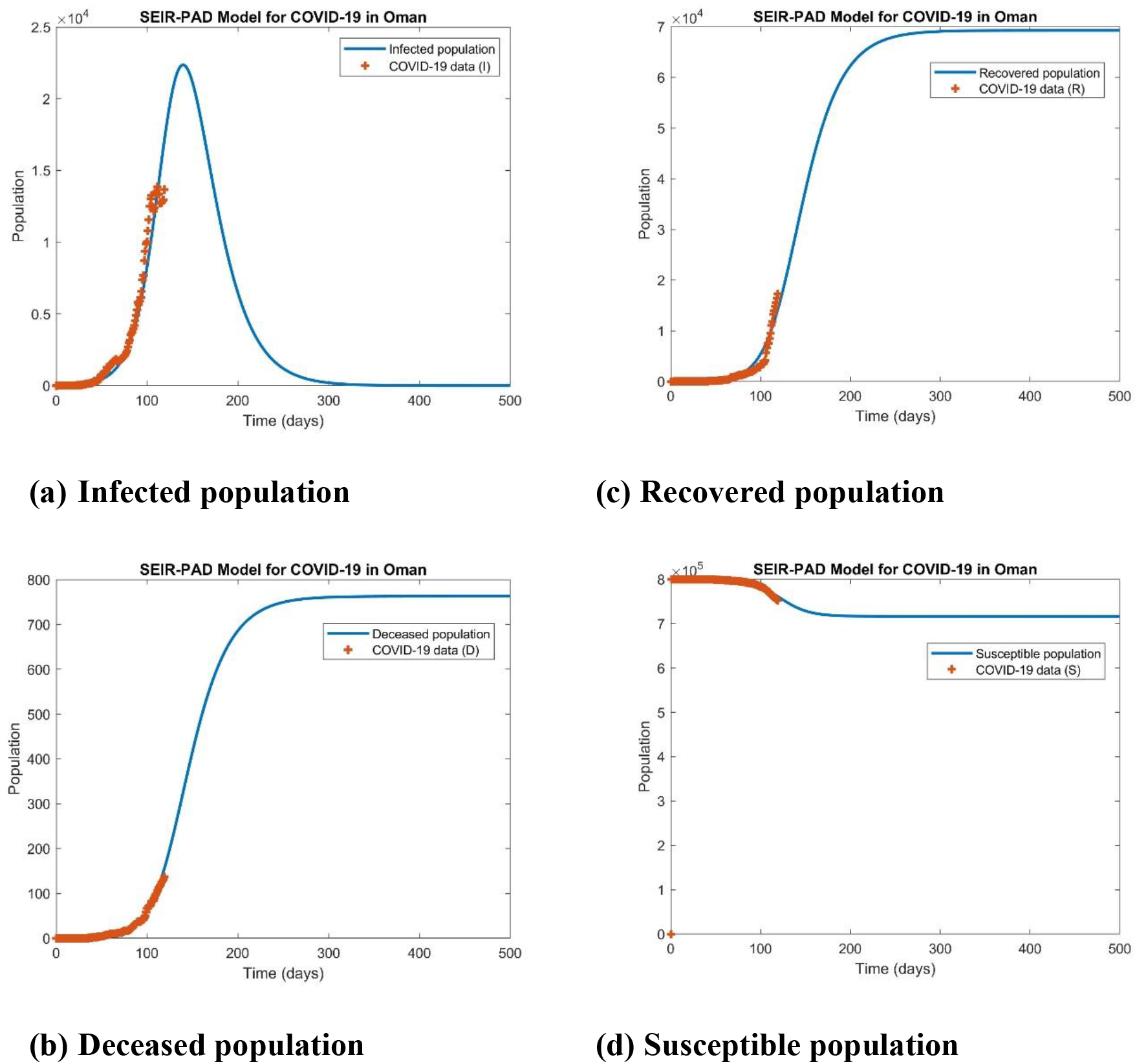
Fitting 4 set of COVID-19 data simultaneously with SEIR-PAD model for dynamics of COVID-19 in Oman (23 June 2020).

Figure 5 shows the same results of SEIR-PAD model for Qatar. Peak day of infection is predicted 30^th^ May 2020 with 30,134 active cases whilst actual peak day was 28^th^ May 2020 with 33,896 active cases (see Fig. 5a). Total expected deceased cases are 152 (see Fig. 5b) cases by expected end of the pandemic after 184 days from start of outbreak; i.e. 1^st^ September 2020. This is based on present preventive measures set by the Qatar government. As shown in Fig. 5c, expected total recovered cases by the end of pandemic is predicted 75,700 cases. Total susceptible population will be 1,212,500 by the end of pandemic (see Fig. 5d).

**Figure 5:**
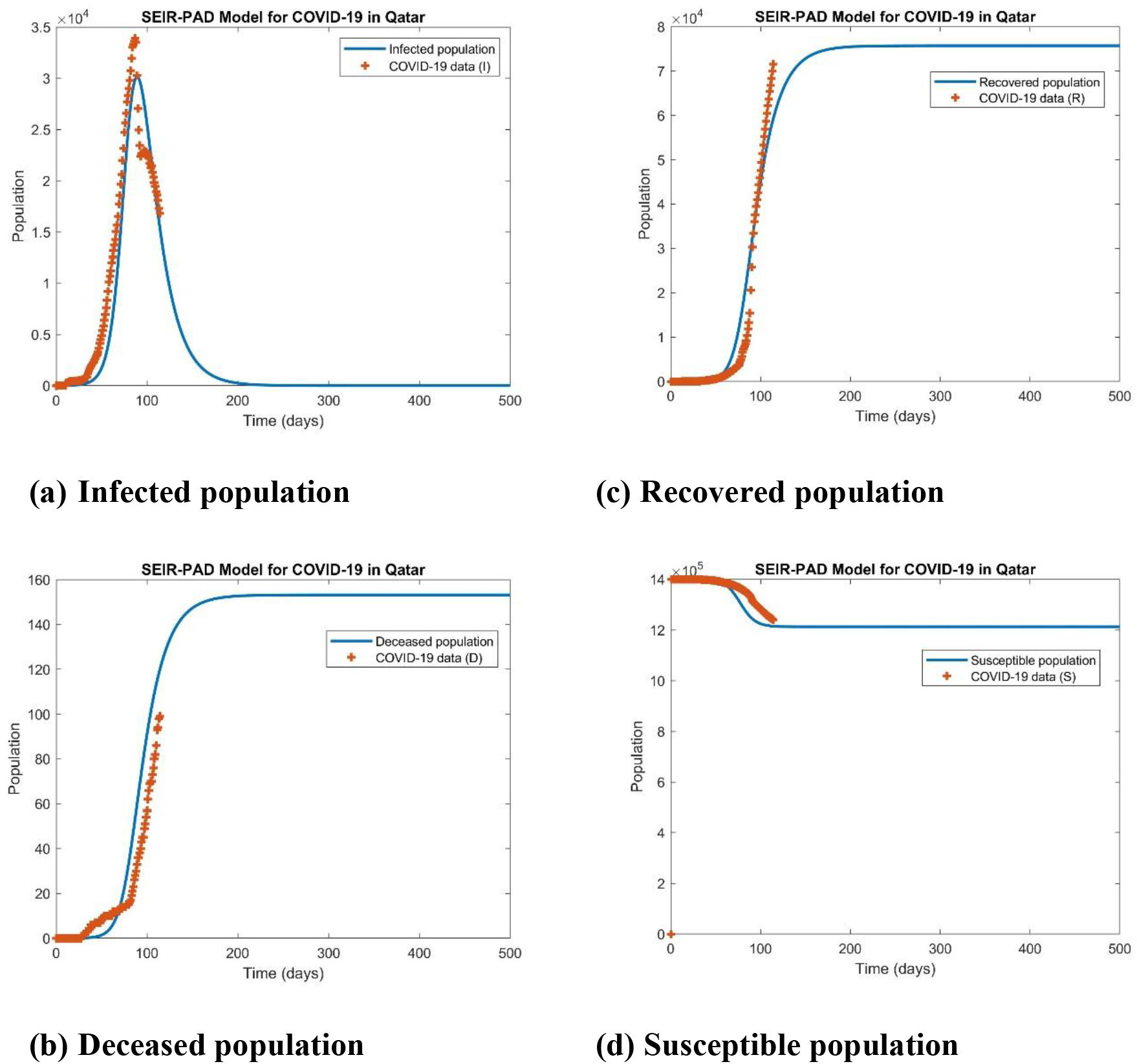
Fitting 4 set of COVID-19 data simultaneously with SEIR-PAD model for dynamics of COVID-19 in Qatar (23 June 2020).

Figure 6 shows the same results of SEIR-PAD model for KSA. Peak day of infection is predicted 6^th^ June 2020 with 39,158 active cases whilst actual peak day was not yet reached on 23^rd^ June 2020 observing a double peak trends with possible second peak on 22^nd^ June 2020 with actual infected population of 51,874 cases (see Fig. 6a). SEIR-PAD model in its present form cannot predict double peak trends. Total expected deceased cases are 1544 cases (see Fig. 6b) by expected end of the pandemic after 176 days from start of outbreak; i.e. 26^th^ August 2020. This is based on present preventive measures set by the KSA government. As shown in Fig. 6c, expected total recovered cases by the end of pandemic is predicted 137,480 cases. Total susceptible population will be 1,338,400 by the end of pandemic (see Fig. 6d).

**Figure 6:**
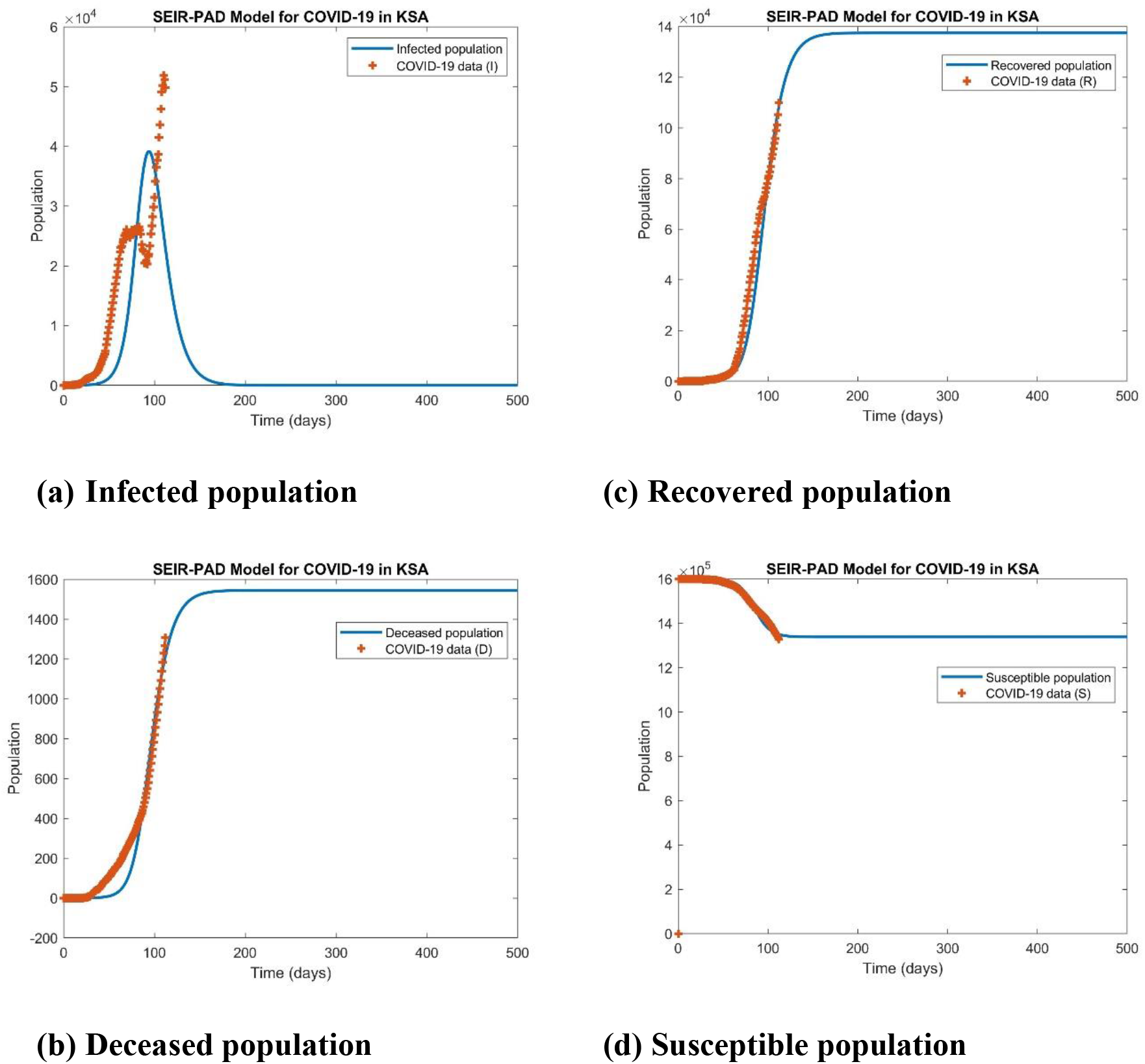
Fitting 4 set of COVID-19 data simultaneously with SEIR-PAD model for dynamics of COVID-19 in KSA (23 June 2020).

Figure 7 shows the same results of SEIR-PAD model for UAE. Peak day of infection is predicted 28^th^ May 2020 with 15,867 active cases whilst actual peak day was 5^th^ June 2020 with 16,517 active cases (see Fig. 7a). Total expected deceased cases are 581 (see Fig. 7b) cases by expected end of the pandemic after 300 days from start of outbreak; i.e. 22^nd^ November 2020. This is based on present preventive measures set by the UAE government. As shown in Fig. 7c, expected total recovered cases by the end of pandemic is predicted 40,580 cases. Total susceptible population will be 51,950 by the end of pandemic (see Fig. 7d).

**Figure 7:**
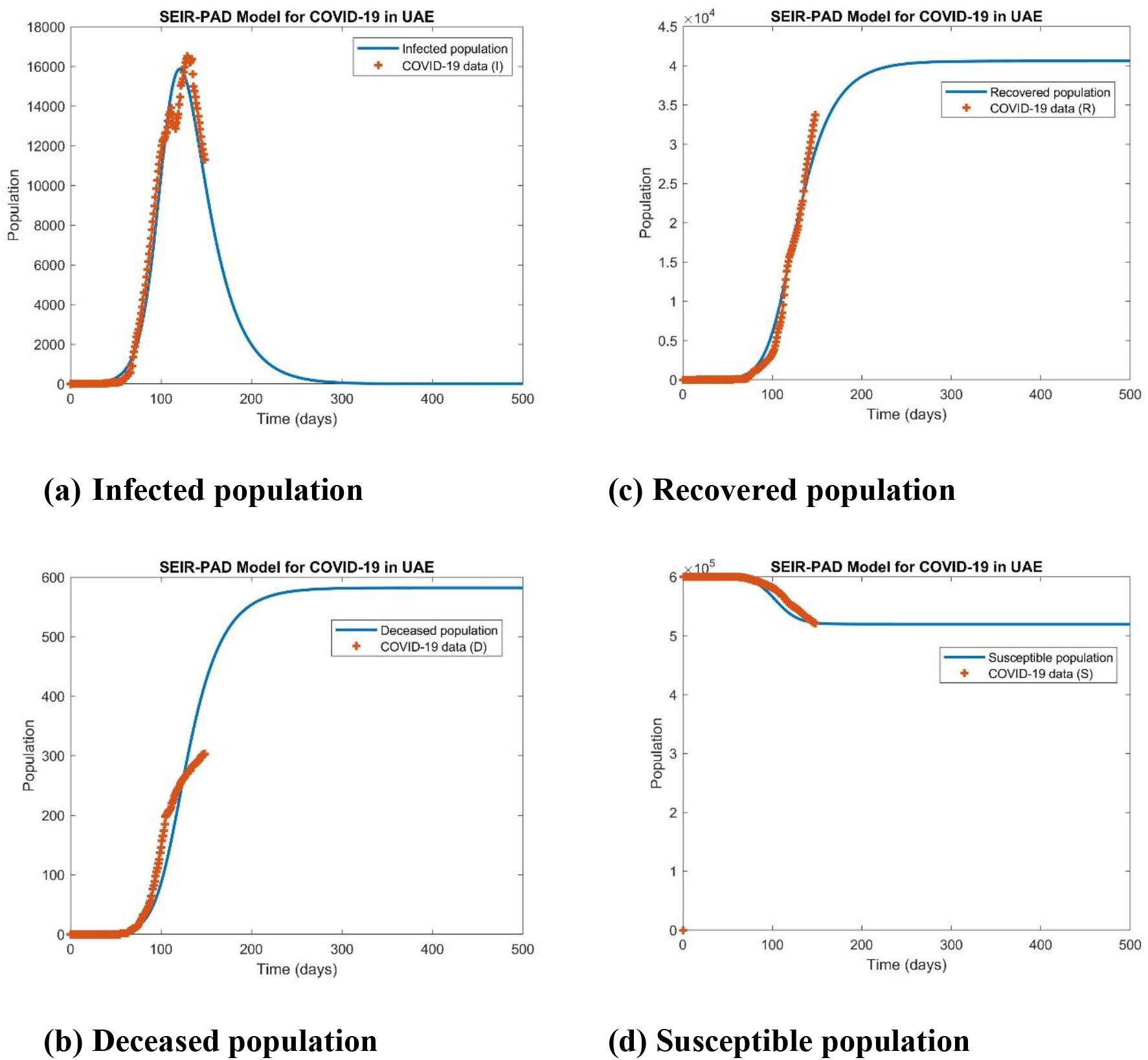
Fitting 4 set of COVID-19 data simultaneously with SEIR-PAD model for dynamics of COVID-19 in UAE (23 June 2020).

## Conclusions

An eight parameter SEIR-PAD model is developed to study the COVID-19 pandemic. The SEIR-PAD is successfully implemented for four populations with available clinical data in MATLAB and best fitted model parameters are obtained using the optimization algorithm discussed here. The SEIR-PAD model is validated for predicting trends of COVID-19 in GCC countries. It is concluded that:

- Peak day of infection is predicted on 13^th^ June 2020 with 12,547 active cases, and the total expected deceased cases of 567 by expected end of the pandemic on 1^st^ October 2020 in Kuwait.
- Peak day of infection is predicted on 16^th^ June 2020 with 5,766 active cases, and the total expected deceased cases of 73 by expected end of the pandemic on 11^th^ September 2020 in Bahrain.
- Peak day of infection is predicted on 15^th^ July 2020 with 22,357 active cases, and the total expected deceased cases of 760 by expected end of the pandemic on 21^st^ December 2020 in Oman.
- Peak day of infection is predicted on 30^th^ May 2020 with 30,134 active cases, and the total expected deceased cases of 152 by expected end of the pandemic on 1^st^ September 2020 in Qatar.
- Peak day of infection is predicted on 6^th^ June 2020 with 39,158 active cases, and the total expected deceased cases of 1544 by expected end of the pandemic on 26^th^ August 2020 in KSA. KSA was the only country in GCC with expected double peak of infectious.
- Peak day of infection is predicted on 28^th^ May 2020 with 15,867 active cases, and the total expected deceased cases of 581 by expected end of the pandemic on 22^nd^ November 2020 in UAE.

SEIR-PADC model is a mathematical model with certain simplified assumptions for better understanding of COVID-19 trends and mechanisms in different countries. The results and outcome of present study depend on the current policies and preventive measures set by the governments or health organizations.

## Data Availability

data is public

https://www.worldometers.info/coronavirus/

